# Utilizing Intraindividual Cognitive Variability to Predict Early Neuronal Synuclein Disease Progression

**DOI:** 10.1101/2025.10.21.25338482

**Authors:** Hannah L. Combs, Ryan Kurth, Anuprita Nair, Michele K. York, Daniel Weintraub, David-Erick Lafontant, Chelsea Caspell-Garcia, the Parkinson’s Progression Markers Initiative

**Author notes:** **Corresponding Author:** Hannah L. Combs, PhD, ABPP, 7200 Cambridge St. 9^th^ Floor, Houston, TX 77030.

## Abstract

**Background:** Neuronal synuclein disease (NSD) involves pathological α-synuclein presence and often dopaminergic dysfunction, initially preceding overt clinical symptoms. NSD-ISS identifies Stage 2A (no dopaminergic dysfunction) and 2B (dopaminergic dysfunction) as prodromal phases marked by subtle clinical signs without functional impairment. Intraindividual variability/ dispersion (IIV-D), reflecting within-person inconsistency across cognitive tasks, has emerged as a potential marker of early neurodegenerative changes.

**Objectives:** This study examined whether IIV-D differentiates NSD Stage 2 participants from healthy controls and predicts progression to more advanced NSD stages.

**Methods:** Data from the Parkinson’s Progression Markers Initiative were used to assess performance across 11 neuropsychological tests in 934 participants (832 Stage 2; 102 controls). IIV-D was quantified using the total coefficient of variation (CoV) and a domain-specific attention/executive CoV. Group comparisons and logistic regression assessed associations between IIV-D, clinical characteristics, and disease progression.

**Results:** Stage 2 participants exhibited significantly greater CoV than controls (*p* = .003). Higher IIV-D was associated with worse motor symptoms, non-motor burden, and functional impairment. Among Stage 2 participants, subsequent converters to Stage 3+ (n = 100) had significantly higher total CoV (*p* = .008) and attention/executive CoV (*p* = .020) at baseline. CoV independently predicted conversion after one year (OR = 1.44, *p* = .008), controlling for baseline motor severity.

**Conclusions:** IIV-D, particularly CoV, may be a sensitive cognitive marker of early NSD and predict short-term disease progression. Findings support integrating cognitive dispersion metrics into early detection strategies for prodromal synucleinopathies, though replication is needed to confirm generalizability and clinical utility.

## INTRODUCTION

The pathogenic processes of Parkinson’s disease (PD) are thought to precede the manifestation of clinical symptoms by years. Identifying and characterizing clinical markers associated with this preclinical stage is vital. In 2024, a new research biological definition and staging system for neuronal α-synuclein disease (NSD-ISS) was proposed^1^. NSD-ISS is defined by the presence of pathological neuronal α-synuclein as the primary biological anchor and with dopaminergic neuronal dysfunction as the secondary biological anchor. The NSD-ISS incorporates an integrated staging system which is based on these biological anchors and the severity of functional impairment caused by clinical signs or symptoms. The presence of meaningful clinical signs marks the transition to NSD-ISS stage 2 and beyond. Stage 2 is characterized by presence of NSD plus subtle signs or symptoms (e.g., hyposmia, iRBD, or early motor or cognitive symptoms) without functional impairment, indicating a prodromal disease state. Stage 2A is without dopaminergic neuronal dysfunction (negative), while Stage 2B has dopaminergic neuronal dysfunction (positive) (1). In the Parkinson’s Progression Markers Initiative (PPMI), the presence of NSD is currently determined by a positive cerebrospinal fluid (CSF) α-synuclein seed amplification assay (SAA), and dopaminergic neuronal dysfunction is assessed using a single-photon emission computerized tomography (SPECT) dopamine transporter (DAT) scan (DaTscan).

Intraindividual cognitive variability (IIV-D), also known as cognitive dispersion, quantifies inconsistency in an individual’s performance across various cognitive tests. Elevated IIV-D across a battery of neuropsychological tests, or dispersion within a neuropsychological profile, has increasingly been linked to breakdowns in neuronal integrity and may reflect diminished top-down executive control over time. This is supported by neuroimaging studies showing associations between IIV-D and brain structures integral to executive networks^2–4^, as well as behavioral research demonstrating elevated IIV-D in individuals with frontal systems injury^5, 6^. IIV-D has also been shown to predict executive dysfunction in daily life ^7^ and is uniquely associated with executive functioning in neuropsychological conditions prone to executive deficits^8^. Increased IIV-D has been observed in clinical populations with known executive dysfunction, including PD^9^ and dementia with Lewy bodies^10^. In a study by Jones and colleagues^9^, increased cognitive dispersion was found to predict cognitive decline in PD, but not healthy controls (HCs). Specifically, greater IIV-D in cognitive performance was associated with higher risk of transitioning from cognitively intact to mild cognitive impairment (MCI). Combs et al.^11^ found greater cognitive dispersion in patients with PD compared with controls, even in individuals classified as having normal cognition. Other studies have suggested IIV-D may also predict functional status^12^ and serve as a marker of longitudinal cortical volume loss^13^. While the relationship between IIV-D and the NSD-ISS has not yet been explored, understanding how IIV-D correlates with NSD stages could provide valuable insights into early detection and progression of PD. Importantly, because cognitive variability may reflect diffuse neuropathological changes, IIV-D could serve as a marker of broader disease progression, not just cognitive decline, highlighting its potential utility in tracking the widespread nature of early synucleinopathy.

The present study utilized the Parkinson’s Progression Markers Initiative (PPMI) database to investigate the utility of IIV-D in characterizing and predicting NSD-ISS progression, particularly within the earliest disease states (Stage 2). Specifically, we aimed to:

1. Determine whether baseline IIV-D and domain-specific variability in attention/executive functioning differentiate Stage 2 participants from HCs
2. Assess whether elevated baseline IIV-D is associated with greater motor symptom severity, non-motor burden, and functional impairment
3. Examine whether higher baseline IIV-D predicts progression from Stage 2 to more advanced NSD-ISS stages (Stage 3 or higher) after one year.

## METHODS

### Participants

Data used in the preparation of this article were obtained on January 21, 2025 from the Parkinson’s Progression Markers Initiative (PPMI) database (www.ppmi-info.org/access-data-specimens/download-data), RRID:SCR 006431^14^. For up-to-date information on the study, visit www.ppmi-info.org (see Appendix 1 for PPMI Author List). This analysis was conducted by the PPMI Statistics Core and used actual dates of activity for participants, a restricted data element not available to public users of PPMI data. This analysis used DAT-SPECT and aSynSAA results for participants of the Prodromal Cohort, obtained from PPMI upon request after approval by the PPMI Data Access Committee.

The PPMI study and its enrollment criteria have been previously described in detail^15, 16^. All participants provided informed consent in accordance with institutional review board guidelines. Individuals without a formal diagnosis of PD were enrolled based on risk factors or prodromal indicators of early synucleinopathy, including hyposmia, isolated REM sleep behavior disorder (iRBD), and relevant genetic variants (e.g., *GBA* and *LRRK2*). PD participants met inclusion criteria based on clinical presentation (e.g., asymmetric resting tremor or bradykinesia, or at least two of resting tremor, bradykinesia, and rigidity), a recent PD diagnosis made by the site investigator, absence of PD medication at study entry, and evidence of dopaminergic dysfunction confirmed via DaTscan. All participants were subsequently classified according to their NSD-ISS stage. CSF was collected and assayed for neuronal α-synuclein using the SAA test, and dopaminergic dysfunction was assessed with DaTscan. All participants included in this analysis were required to have completed all eight neuropsychological tests from the expanded PPMI cognitive battery used to calculate IIV-D (described below). Since some participants entered the study before the implementation of the expanded battery, “baseline” was defined for the purposes of this study as the first visit in which a participant had completed all eight tests of the battery. The Stage 2 cohort included only participants who were classified as Stage 2 (defined by subtle signs or symptoms without functional impairment, or prodromal disease) at this baseline visit. Participants classified as Stage 3 (signs or symptoms with slight functional impairment) and above were excluded. Stage 2A and 2B designations were based on DaTscan results, with Stage 2B defined by abnormal DAT binding (<75% of age- and sex-adjusted lowest putamen specific binding ratio).

HCs were required to have no clinically significant neurological dysfunction, no first-degree relative with PD, and a baseline Montreal Cognitive Assessment (MoCA) score ≥27. For inclusion in the present analyses, HCs also could not exhibit hyposmia and were required to have a negative CSF neuronal α-synuclein SAA test. To exclude HCs with incipient cognitive decline, this group was further required to have a Year 1 MoCA score ≥26 and no more than a 2-point decline between baseline and Year 1 assessments.

### Measures

The following neuropsychological measures were administered as part of the PPMI expanded cognitive battery and used in the present study to examine group differences and to calculate IIV-D: Benton Judgement of Line Orientation (JLO)^17^, Hopkins Verbal Learning Test-Revised (HVLT-R Total Recall, Delayed Recall, & Recognition Trials)^18^, Wechsler Adult Intelligence Scale, 4^th^ Edition Letter- Number Sequencing (WAIS-IV LNS)^19^, Controlled Oral Word Association Test (COWAT)^20^, Modified Boston Naming Test (BNT), modified semantic fluency ^21^, Symbol Digit Modalities Test (SDMT)^22^, and Trail Making Test (TMT) Parts A & B ^20^. The following motor and non-motor assessments were included to examine correlates of IIV-D: Movement Disorder Society Unified Parkinson’s Disease Rating Scale (MDS-UPDRS)^23^, Scales for Outcomes in Parkinson’s disease- Autonomic Dysfunction (SCOPA-AUT)^24^, Geriatric Depression Scale, 15 item (GDS-15)^25^, Questionnaire for Impulsive-Compulsive Disorders in Parkinson’s Disease (QUIP)^26^, State-Trait Anxiety Inventory, State Subscale (STAI State), University of Pennsylvania Smell Identification Test (UPSIT)^27^, Epworth Sleepiness Scale (ESS)^28^, the REM Sleep Behavior Disorder Questionnaire (RBDSQ)^29^ and the Montreal Cognitive Assessment (MoCA)^30^.

### IIV-D Dispersion

All individual test scores on the neuropsychological measures were corrected based on internally-derived normative data and converted to a common metric, a *T* score, with a mean of 50 and standard deviation of 10. Internal norms were developed using linear regression models that included age, sex, and education. However, demographic corrections were applied selectively, depending on whether these variables significantly affected performance on each test. Details regarding this normative process have been described previously^31^. A composite score reflecting average cognitive performance was established by calculating the mean of all 11 neurocognitive test scores. IIV-D can be measured in many ways and in the present study was examined based on the coefficient of variance (CoV). CoV has been used to examine IIV-D in multiple populations^8, 32–35^. The CoV was calculated by dividing the standard deviation of a participant’s 11 normed test scores, computed as the square root of the average squared deviation from the mean, by their mean score across those tests’ (mean composite score)^36^. The Attention/Executive CoV was calculated in the same manner, restricting the calculation to the five PPMI internal normatively derived attention/executive scores (Symbol Digit Modalities Test Oral Total, Trail Making Test Part A Total, Letter-Number Sequencing Total, Lexical Fluency Total, & Trail Making Test Part B). Higher CoV indicates greater dispersion in performance, which reflects poorer cognitive consistency.

### Data Analysis

Statistical analyses were performed using SAS v9.4 (SAS Institute Inc., Cary, NC; sas.com; RRID:SCR_008567). Statistical analysis codes used to perform the analyses in this article are shared on Zenodo [10.5281/zenodo.17252636].

Summary statistics were calculated at baseline for all variables stratified by NSD Stage 2 and non-NSD HC groups. Chi-square tests were run for the categorical demographic variables, and two sample t-tests were utilized for the continuous variables, both demographic and variables of interest. Welch’s t-test was used in cases where the equality of variance assumption was unmet (an F-test with p <0.05). For each comparison, the t-value, p-value, and Cohen’s d effect size are reported. This procedure was repeated on just the Stage 2 cohort, stratified by conversion vs. non-conversion to NSD Stage 3 or higher after one year. Only those who had completed the expanded cognitive battery for at least two visits were included in this second round of analyses. To account for multiple comparisons and control the false discovery rate (FDR), we applied the Benjamini-Hochberg (BH) procedure to the list of p-values obtained from our statistical tests. This method ranks p-values in ascending order and compares each to a calculated threshold based on its rank, the total number of tests, and the desired FDR level (set at 0.05). P-values that met the BH-adjusted significance threshold were considered statistically significant. This approach balances the risk of Type I errors while maintaining statistical power, making it appropriate for exploratory analyses involving multiple hypotheses. Associations between IIV-D metrics (total CoV and Attention/Executive CoV) and clinical characteristics, including motor symptoms, non-motor burden, and functional scores were examined via Pearson correlation coefficients (r). To assess progression of NSD staging over time and whether greater IIV-D predicts conversion to a higher stage, binomial logistic regression models were utilized.

The first model included conversion from NSD Stage 2 to Stage 3 or higher after one year as the binary outcome variable with baseline CoV as predictor variable, controlling for baseline motor severity (MDS-UPDRS Part III score). The second model included attention/executive CoV at baseline as the predictor variable.

## RESULTS

### Patient Characteristics

A total of 934 participants were included in the analyses, comprising 832 individuals classified as NSD-ISS Stage 2 and 102 HCs. Demographic comparisons revealed that Stage 2 participants were significantly older than controls (*M* = 67.77 vs. 64.64 years; *t* = −2.56, *p* = .012), but groups did not differ significantly in education, race/ethnicity, or sex (see Table 1). Clinically, Stage 2 participants exhibited significantly greater motor symptom severity as measured by MDS-UPDRS Part III score (*p* < 0.001) and Hoehn & Yahr stage (*p* < 0.001), as well as higher scores on measures of autonomic dysfunction (SCOPA-AUT), depression (GDS-15), anxiety (STAI State), and sleep disturbance (ESS and RBDSQ). They also demonstrated significantly lower olfactory function (UPSIT) and reduced performance across MoCA and multiple cognitive domains, including verbal memory, fluency, and processing speed. Notably, despite these group differences, cognitive performance among Stage 2 participants remained within normal limits. The mean composite T score for this group was 47, which falls within the average range (T score mean = 50, SD = 10), indicating that NSD Stage 2 participants were, on average, cognitively intact at baseline.

**Table 1.**
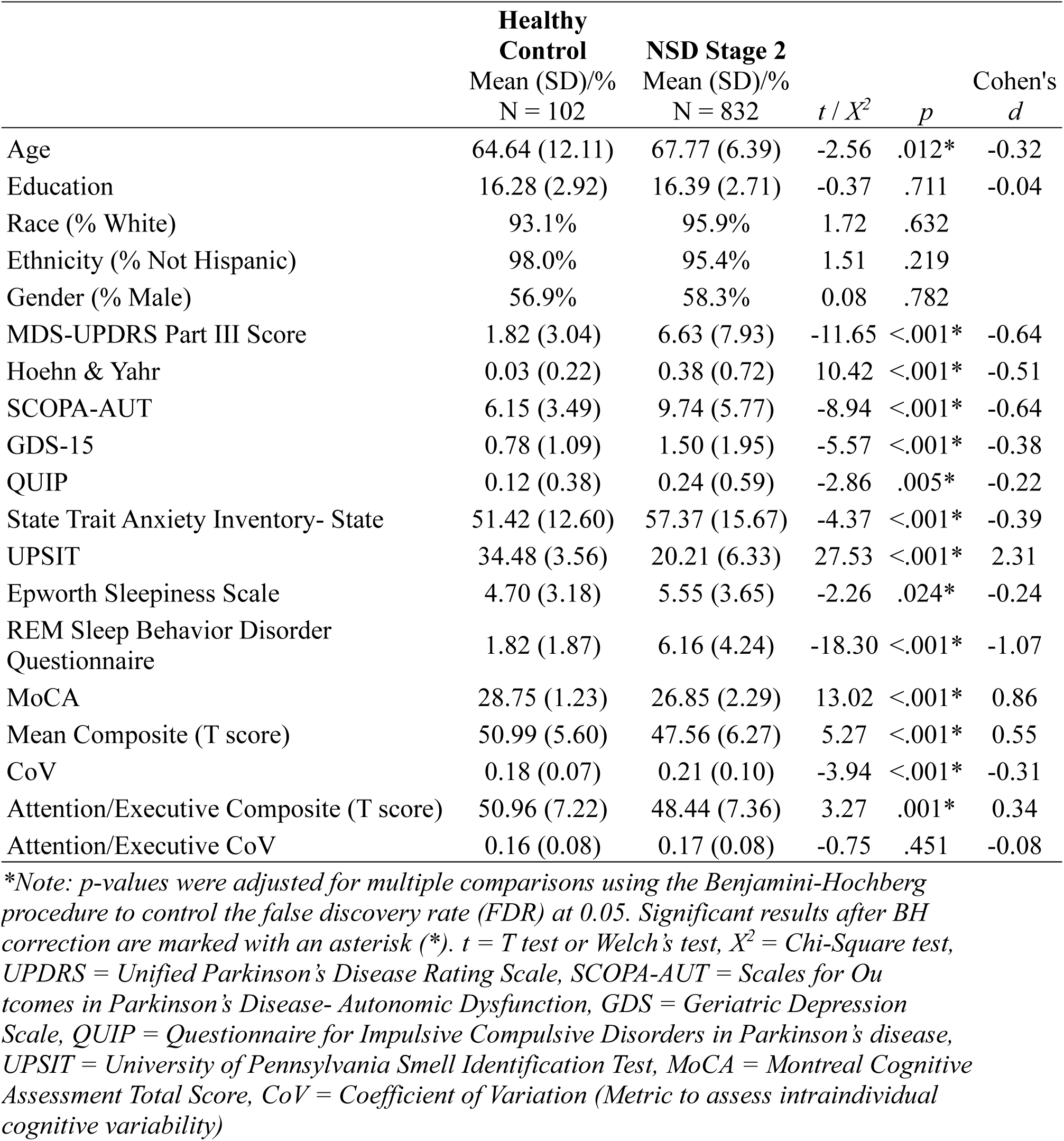
NSD Stage 2 and Healthy Control Group Comparisons.

NSD Stage 2 demonstrated significantly greater total CoV compared with HCs (*t*(932) = −3.94, *p* < .001, d = −0.31, see Table 1). No significant group differences were observed for attention/executive CoV. Across the full cohort (i.e., Stage 2 and HCs), total CoV was positively correlated with age (*r* = 0.124, *p* < .001), UPDRS Part III score (*r* = .094, *p* = .005), GDS-15 (*r* = .116, *p* < .001), STAI State (*r* = .155, *p* < .001), and negatively correlated with education (*r* = −.192, *p* < .001) and UPSIT (r = −.117, *p* = .001). Attention/executive CoV was not significantly correlated with other motor or non-motor measures. Within the Stage 2 group, total CoV remained significantly associated with age (r = .135, *p* < .001), education (r = −.201, *p* < .001), GDS (*r=* .107, *p* = .002), and STAI (r = .154, *p* < .001). Attention/executive CoV was significantly correlated with STAI (r = .091, *p* = .009).

### Progression in NSD-ISS Staging

Figure 1 illustrates one-year NSD-ISS stage transitions among participants initially classified as NSD Stage 2. Among Stage 2 participants, those who converted to NSD-ISS Stage 3 or higher after one year (n = 100) demonstrated significantly greater baseline total CoV (*p* = .008, d = −0.30) and attention/executive CoV (*p* = .020, d = −0.27). They also demonstrated significantly greater baseline motor symptom severity compared to those who did not convert (n = 340). Specifically, converters had higher scores on UPDRS Part III (*p* < .001, d = −0.80) and Hoehn & Yahr staging (*p* < .001, d = −0.77). Cognitive performance was also lower among converters, with significantly reduced overall mean composite score (*p* < .001, d = 0.46) and attention/executive composite score (*p* < .001, d =0.50). See Table 2 for comparisons of converters and non-converters after year one.

**Figure 1.**
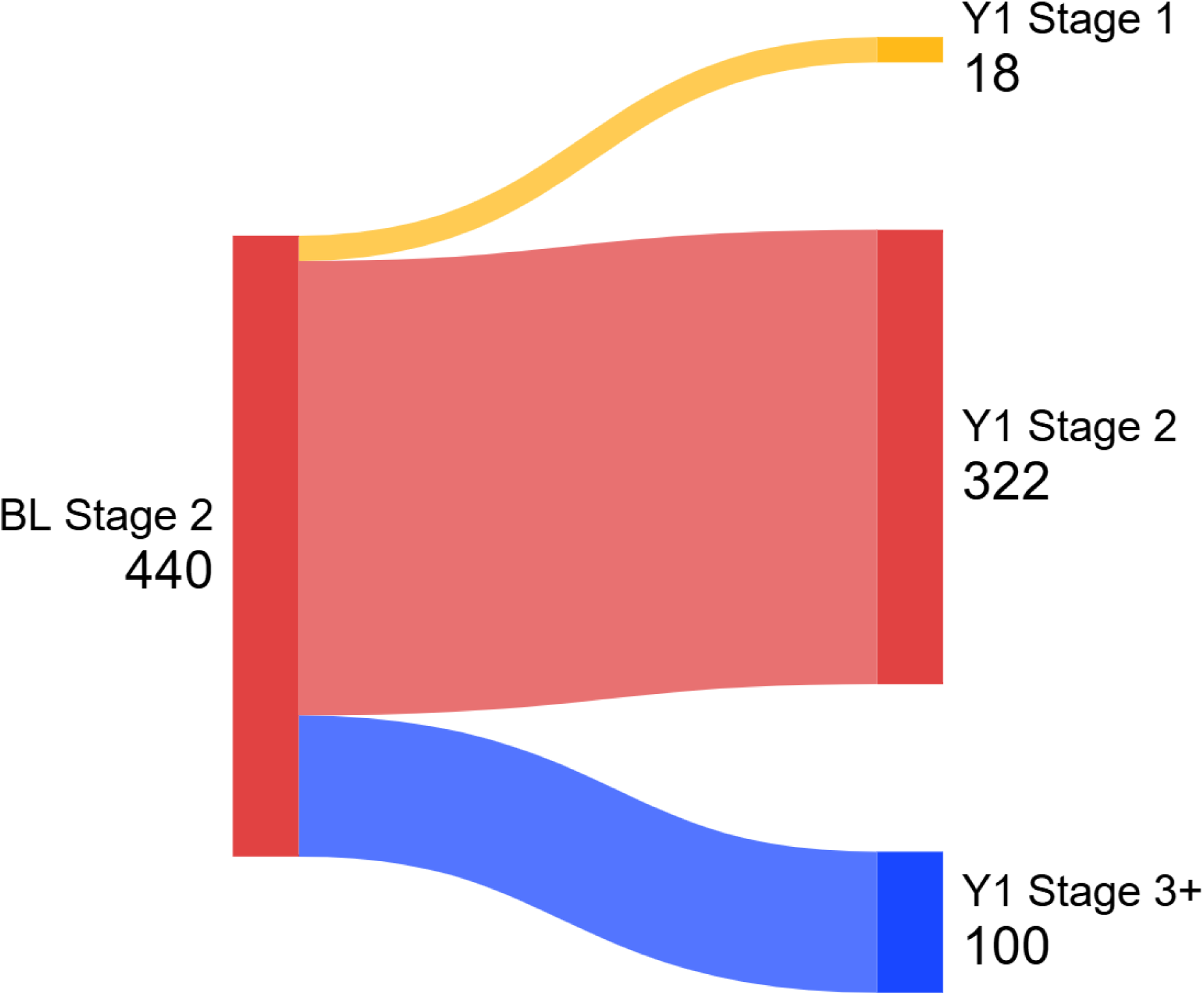
One-Year Progression Among NSD Stage 2 Participants. *\*Note: Sankey diagram showing transitions of Stage 2 participants from baseline to Year 1. The flow widths are proportional to the number of participants*.

**Table 2.** Group Comparisons of NSD Stage 2 Converters after Year 1.

|  | **Did not**<br>**Convert**<br>Mean (SD)/%<br>N = 340 | **Converted to**<br>**Stage 3+**<br>Mean (SD)/%<br>N = 100 | *t* | *p* | Cohen's *d* |
| --- | --- | --- | --- | --- | --- |
| Mean Composite (T score) | 48.53 (6.22) | 45.66 (6.28) | 4.04 | <0.001* | 0.46 |
| CoV | 0.19 (0.08) | 0.22 (0.08) | -2.66 | 0.008* | -0.30 |
| Attention/Executive Composite | 49.62 (7.37) | 45.93 (7.60) | 4.37 | <0.001* | 0.50 |
| Attention/Executive CoV | 0.16 (0.08) | 0.18 (0.07) | -2.58 | 0.011* | -0.27 |
| Age | 67.52 (6.10) | 67.74 (7.51) | -0.26 | 0.0792 | -0.03 |
| Education | 16.26 (2.80) | 16.49 (2.68) | -0.72 | 0.471 | -0.08 |
| MDS-UPDRS Part III | 5.61 (6.69) | 11.73 (10.21) | -5.60 | <0.001* | -0.80 |
| Hoehn & Yahr | 0.28 (0.63) | 0.82 (0.88) | -5.59 | <0.001* | -0.77 |
| SCOPA-AUT | 9.58 (5.63) | 11.00 (6.91) | -1.87 | 0.063 | -0.24 |
| GDS-15 | 1.60 (2.16) | 1.77 (2.30) | -0.69 | 0.491 | -0.08 |
| QUIP | 0.26 (0.62) | 0.32 (0.68) | -0.80 | 0.426 | -0.09 |
| State Trait Anxiety Inventory- State | 57.42 (16.67) | 61.05 (18.18) | -1.87 | 0.062 | -0.21 |
| UPSIT | 20.77 (6.30) | 20.87 (6.76) | -0.13 | 0.900 | -0.02 |
| Epworth Sleepiness Scale | 5.70 (3.60) | 6.39 (4.29) | -1.47 | 0.145 | -0.18 |
| REM Sleep Behavior Disorder<br>Questionnaire | 6.18 (4.35) | 5.87 (4.22) | 0.64 | 0.524 | 0.07 |

*\*Note: p-values were adjusted for multiple comparisons using the Benjamini-Hochberg procedure to control the false discovery rate (FDR) at 0.05. Significant results after BH correction are marked with an asterisk (\*). t = T test or Welch's test, CoV = Coefficient of Variation (Metric to assess intraindividual cognitive variability), UPDRS = Unified Parkinson's Disease Rating Scale, SCOPA-AUT = Scales for Outcomes in Parkinson's Disease- Autonomic Dysfunction, GDS = Geriatric Depression Scale, QUIP = Questionnaire for Impulsive Compulsive Disorders in Parkinson's disease, UPSIT = University of Pennsylvania Smell Identification Test*

Two binary logistic regressions were conducted to examine whether IIV-D predicted conversion from NSD Stage 2 to Stage 3 or higher after one year, while controlling for motor severity (UPDRS Part III score). In the first model, the overall combination of predictors (total CoV and UPDRS Part III) significantly improved model fit compared to a null model (χ²(2) = 46.08, *p* < .001). The first model demonstrated acceptable fit (Hosmer-Lemeshow test: χ² = 5.03, *p* = .755) and explained approximately 15% of the variance in conversion status (Nagelkerke R² = .154). Individually, total CoV was a significant independent predictor of conversion after one year (*B* = 0.362, *p* = .008), with an odds ratio of 1.44 (95% CI: 1.10-1.88), suggesting that higher CoV was associated with increased odds of conversion. UPDRS Part III score also significantly predicted conversion (*B* = 0.09, *p* < .001), with each point increase associated with an 8.6% increase in the odds of conversion (95% CI: 1.06–1.12). The second model (attention/executive CoV and UPDRS Part III) was also significant (χ²(2) = 44.35, *p* < .001), explaining 15% of the variance in conversion status (Nagelkerke R² = .149) with acceptable model fit (Hosmer-Lemeshow test: χ² = 13.90, *p* = .084). Attention/executive CoV was a significant independent predictor of conversion after one year (*B* = 0.324, *p* = .023), with an odds ratio of 1.38 (95% CI: 1.05-1.83), suggesting that higher attention/executive CoV was also associated with increased odds of conversion.

## DISCUSSION

The present study examined IIV-D as a potential marker of early NSD and its progression using data from PPMI. Our findings support the utility of IIV-D, particularly the CoV, in differentiating individuals at NSD-ISS Stage 2 from HC and in predicting progression to more advanced disease stages after one year. Consistent with prior literature on cognitive dispersion in neurodegenerative conditions, Stage 2 participants demonstrated significantly greater IIV-D compared with HCs, even in the absence of overt cognitive impairment. This suggests that cognitive inconsistency may emerge early in the disease process, potentially reflecting subtle disruptions in neural integrity. Notably, while overall CoV was elevated in Stage 2, domain-specific variability in attention/executive functioning did not differ significantly between groups, highlighting the importance of broad cognitive dispersion metrics in early detection. Correlational analyses revealed that higher IIV-D was associated with worse motor symptoms, greater non-motor burden, and lower functional status. These findings align with emerging evidence that cognitive variability may reflect diffuse frontal systems network changes, including motor and autonomic systems^13^.

The present study also highlights the potential role of CoV in detecting disease progression. Among individuals initially classified as Stage 2, those who converted to Stage 3 or higher after one year demonstrated significantly greater IIV-D as measured by both overall CoV and attention/executive CoV. These results suggest that elevated IIV-D may serve as an early indicator of disease progression, consistent with prior work linking cognitive instability to neurodegenerative trajectories^13^. Importantly, logistic regression analyses revealed that total CoV remained a significant independent predictor of conversion even after controlling motor severity. The odds ratio associated with CoV (OR = 1.41) suggested that individuals with greater variability in cognitive performance were approximately 41% more likely to progress to a higher disease stage, compared to those with lower variability. Attention/executive CoV did not reach statistical significance in follow-up models.

Although mean cognitive performance was significantly lower among Stage 2 participants and converters, the average t score remained within the normal range, suggesting intact cognition at the group level. However, CoV offers a complementary perspective by capturing intraindividual variability across cognitive domains. This variability may reflect early disruptions in neural networks that are not yet evident in mean-level performance. Clinically, this distinction is important: individuals may appear cognitively intact based on average scores yet exhibit meaningful variability that signals underlying neurodegenerative processes. Incorporating CoV into clinical assessments could enhance early detection and risk stratification by identifying subtle cognitive instability that precedes measurable impairment. As such, CoV may serve as a valuable adjunct to traditional mean-based metrics in both research and clinical settings.

Despite the promising findings, several limitations should be acknowledged. First, the study relied on data from a single cohort (PPMI), which may limit generalizability. Although PPMI is a well-characterized and diverse dataset, replication in independent samples is necessary to confirm the robustness and clinical utility of IIV-D as a predictive marker of NSD-ISS. Second, while the CoV and attention/executive CoV metrics provide valuable insights into cognitive variability, they are influenced by the number and type of neuropsychological tests included. The dispersion metrics used in this study were derived from a fixed battery, and results may differ with alternative test compositions or scoring methods. Additionally, the internal normative corrections applied to test scores, while rigorous, may not fully account for cultural, linguistic, or socioeconomic factors that influence cognitive performance. Third, although longitudinal models demonstrated predictive utility of IIV-D, the follow-up period was relatively short (one year for conversion analyses). Longer-term studies are needed to determine whether IIV-D continues to predict progression over extended periods and whether it can distinguish between different trajectories of cognitive and functional decline. Future analyses should also examine whether IIV-D differs between NSD-ISS Stage 2A and 2B, as the presence of dopaminergic dysfunction may influence cognitive variability.

Taken together, these findings underscore the potential of IIV-D as a non-invasive, scalable biomarker for early NSD detection and monitoring. Cognitive dispersion may reflect underlying pathophysiological processes that precede measurable cognitive impairment, offering a window into disease progression before traditional markers become evident. Future research should explore the neurobiological correlates of IIV-D in NSD, including imaging and fluid biomarkers, and evaluate its utility in clinical trial enrichment and individualized risk stratification.

## Data Availability

Data used in the preparation of this article were obtained on January 21, 2025 from the Parkinsons Progression Markers Initiative (PPMI) database (www.ppmi-info.org/access-data-specimens/download-data), RRID:SCR 006431. This analysis was conducted by the PPMI Statistics Core and used actual dates of activity for participants, a restricted data element not available to public users of PPMI data. This analysis used DAT-SPECT and aSynSAA results for participants of the Prodromal Cohort, obtained from PPMI upon request after approval by the PPMI Data Access Committee. Statistical analysis codes used to perform the analyses in this article are shared on Zenodo [10.5281/zenodo.17252636].

## Financial Disclosure/Conflict of Interest

H.C. has received research funding from The Michael J. Fox Foundation for Parkinson’s Research. D.W. has received research funding or support from The Michael J. Fox Foundation for Parkinson’s Research, International Parkinson and Movement Disorder Society, National Institutes of Health, The Parkinson’s Foundation, and the US Department of Veterans Affairs; honoraria for consultancy from Boehringer Ingelheim, Cerevel Therapeutics, CHDI Foundation, Citrus Health Group, Medscape, Modality.AI, Roche, Sage Therapeutics, Scion NeuroStim, and Signant Health; and license fee payments from the University of Pennsylvania for the Questionnaire for Impulsive-Compulsive Disorders in Parkinson’s Disease (QUIP) and Questionnaire for Impulsive Compulsive Disorders in Parkinson’s Disease-Rating Scale (QUIPRS). M.K.Y. has received research funding or support from the Alzheimer’s Association, the Michael J. Fox Foundation for Parkinson’s Research, the Parkinson Foundation, and BlueRock Therapeutics. R.K., A.N., D.E.L, and C.C.G. report research funding to their institution from the Michael J. Fox Foundation for Parkinson’s Research.

## Funding Sources for Study

This work was supported by The Michael J. Fox Foundation for Parkinson’s Research (Michael J. Fox Foundation Write Now Initiative Award).

## Acknowledgements

Statistical analysis codes used to perform the analyses in this article are shared on Zenodo [10.5281/zenodo.17252636]. PPMI – a public-private partnership – is funded by the Michael J. Fox Foundation for Parkinson’s Research and funding partners, including 4D Pharma, Abbvie, AcureX, Allergan, Amathus Therapeutics, Aligning Science Across Parkinson’s, AskBio, Avid Radiopharmaceuticals, BIAL, BioArctic, Biogen, Biohaven, BioLegend, BlueRock Therapeutics, Bristol-Myers Squibb, Calico Labs, Capsida Biotherapeutics, Celgene, Cerevel Therapeutics, Coave Therapeutics, DaCapo Brainscience, Denali, Edmond J. Safra Foundation, Eli Lilly, Gain Therapeutics, GE HealthCare, Genentech, GSK, Golub Capital, Handl Therapeutics, Insitro, Jazz Pharmaceuticals, Johnson & Johnson Innovative Medicine, Lundbeck, Merck, Meso Scale Discovery, Mission Therapeutics, Neurocrine Biosciences, Neuron23, Neuropore, Pfizer, Piramal, Prevail Therapeutics, Roche, Sanofi, Servier, Sun Pharma Advanced Research Company, Takeda, Teva, UCB, Vanqua Bio, Verily, Voyager Therapeutics, the Weston Family Foundation and Yumanity Therapeutics

## Appendix 1. PPMI STUDY TEAMS/CORES/COLLABORATORS FOR PUBLICATIONS

### Executive Steering Committee

Kenneth Marek, MD^1^ (Principal Investigator); Caroline Tanner, MD, PhD^9^; Tanya Simuni, MD^3^; Andrew Siderowf, MD, MSCE^12^; Douglas Galasko, MD^27^; Lana Chahine, MD^41^; Christopher Coffey, PhD^4^; Kalpana Merchant, PhD^61^; Kathleen Poston, MD^40^; Roseanne Dobkin, PhD^43^; Tatiana Foroud, PhD^15^; Brit Mollenhauer, MD^8^; Dan Weintraub, MD^12^; Ethan Brown, MD^9^; Karl Kieburtz, MD, MPH^23^; Mark Frasier, PhD^6^; Todd Sherer, PhD^6^; Sohini Chowdhury, MA^6^; Roy Alcalay, MD^36^ and Aleksandar Videnovic, MD^47^

### Steering Committee

Duygu Tosun-Turgut, PhD^9^; Werner Poewe, MD^7^; Susan Bressman, MD^14^; Jan Hammer^15^; Raymond James, RN^22^; Ekemini Riley, PhD^42^; John Seibyl, MD^1^; Leslie Shaw, PhD^12^; David Standaert, MD, PhD^18^; Sneha Mantri, MD, MS^62^; Nabila Dahodwala, MD^12^; Michael Schwarzschild^47^; Connie Marras^45^; Hubert Fernandez, MD^25^; Ira Shoulson, MD^23^; Helen Rowbotham^2^; Paola Casalin^11^ and Claudia Trenkwalder, MD^8^

### Michael J. Fox Foundation (Sponsor)

Todd Sherer, PhD; Sohini Chowdhury, MA; Mark Frasier, PhD; Jamie Eberling, PhD; Katie Kopil, PhD; Alyssa O’Grady; Maggie McGuire Kuhl; Leslie Kirsch, EdD and Tawny Willson, MBS

### Study Cores, Committees and Related Studies

*Project Management Core:* Emily Flagg, BA^1^

*Site Management Core:* Tanya Simuni, MD^3^; Bridget McMahon, BS^1^

*Strategy and Technical Operations:* Craig Stanley, PhD^1^; Kim Fabrizio, BA^1^

*Data Management Core:* Dixie Ecklund, MBA, MSN^4^; Trevis Huff, BSE^4^

*Screening Core:* Tatiana Foroud, PhD^15^; Laura Heathers, BA^15^; Christopher Hobbick, BSCE^15^; Gena Antonopoulos, BSN^15^

*Imaging Core:* John Seibyl, MD^1^; Kathleen Poston, MD^40^

*Statistics Core*: Christopher Coffey, PhD^4^; Chelsea Caspell-Garcia, MS^4^; Michael Brumm, MS^4^

*Bioinformatics Core*: Arthur Toga, PhD^10^; Karen Crawford, MLIS^10^

*Biorepository Core:* Tatiana Foroud, PhD^15^; Jan Hamer, BS^15^

*Biologics Review Committee*: Brit Mollenhauer^8^; Doug Galasko^27^; Kalpana Merchant^61^

*Genetics Core:* Andrew Singleton, PhD^13^

*Pathology Core:* Tatiana Foroud, PhD^15^; Thomas Montine, MD, PhD^40^

*Found:* Caroline Tanner, MD PhD^9^

*PPMI Online:* Carlie Tanner, MD PhD^9^; Ethan Brown, MD^9^; Lana Chahine, MD^41^; Roseann Dobkin, PhD^43^; Monica Korell, MPH^9^

### Site Investigators

Charles Adler, PhD^51^; Roy Alcalay, MD^36^; Amy Amara, PhD^52^; Paolo Barone, PhD^30^; Bastiaan Bloem, PhD^60^ Susan Bressman, MD^14^; Kathrin Brockmann, MD^26^; Norbert Brüggemann, MD^59^; Lana Chahine, MD^41^; Kelvin Chou, MD^44^; Nabila Dahodwala, MD^12^; Alberto Espay, MD^32^; Stewart Factor, DO^16^; Hubert Fernandez, MD^25^; Michelle Fullard, MD^52^; Douglas Galasko, MD^27^; Robert Hauser, MD^19^; Penelope Hogarth, MD^17^; Shu-Ching Hu, PhD^21^; Michele Hu, PhD^58^; Stuart Isaacson, MD^31^; Christine Klein, MD^59^; Rejko Krueger, MD^2^; Mark Lew, MD^49^; Zoltan Mari, MD^56^; Connie Marras, PhD^45^; Maria Jose Martí, PhD^34^; Nikolaus McFarland, PhD^54^; Tiago Mestre, PhD^46^; Brit Mollenhauer, MD^8^; Emile Moukheiber, MD^28^; Alastair Noyce, PhD^63^ Wolfgang Oertel, PhD^64^; Njideka Okubadejo, MD^65^; Sarah O’Shea, MD^39^; Rajesh Pahwa, MD^48^; Nicola Pavese, PhD^57^; Werner Poewe, MD^7^; Ron Postuma, MD^55^; Giulietta Riboldi, MD^53^; Lauren Ruffrage, MS^18^; Javier Ruiz Martinez, PhD^35^; David Russell, PhD^1^; Marie H Saint-Hilaire, MD^22^; Neil Santos, BS^51^; Wesley Schlett^47^; Ruth Schneider, MD^23^; Holly Shill, MD^50^; David Shprecher, DO^24^; Tanya Simuni, MD^3^; David Standaert, PhD^18^; Leonidas Stefanis, PhD^38^; Yen Tai, PhD^29^; Caroline Tanner, PhD^9^; Arjun Tarakad, MD^20^; Eduardo Tolosa PhD^34^ and Aleksandar Videnovic, MD^47^

### Coordinators

Susan Ainscough, BA^30^; Courtney Blair, MA^18^; Erica Botting^19^; Isabella Chung, BS^56^; Kelly Clark^24^; Ioana Croitoru^35^; Kelly DeLano, MS^32^; Iris Egner, PhD^7^; Fahrial Esha, BS^53^; May Eshel^36^; Frank Ferrari, BS^44^; Victoria Kate Foster^57^; Alicia Garrido, MD^34^; Madita Grümmer^59^; Bethzaida Herrera^50^; Ella Hilt^26^; Chloe Huntzinger, BA^52^; Raymond James, BS^22^; Farah Kausar, PhD^9^; Christos Koros, MD, PhD^38^; Yara Krasowski^60^; Dustin Le, BS^17^; Ying Liu, MD^52^; Taina M. Marques, PhD^2^; Helen Mejia Santana, MA^39^; Sherri Mosovsky, MPH^41^; Jennifer Mule, BS^25^; Philip Ng, BS^45^; Lauren O’Brien^48^; Abiola Ogunleye, PGDip^29^; Oluwadamilola Ojo, MD^65^; Obi Onyinanya, BS^28^; Lisbeth Pennente, BA^31^; Romina Perrotti^55^; Michael Pileggi, MS^55^; Ashwini Ramachandran, MSc^12^; Deborah Raymond, MS^14^; Jamil Razzaque, MS^58^; Shawna Reddie, BA^46^; Kori Ribb, BSN,^28^; Kyle Rizer, BA^54^; Janelle Rodriguez, BS^27^; Stephanie Roman, HS^1^; Clarissa Sanchez, MPH^20^; Cristina Simonet, PhD^29^; Anisha Singh, BS^23^; Elisabeth Sittig^64^; Barbara Sommerfeld MSN^16^; Angela Stovall, BS^44^; Bobbie Stubbeman, BS^32^; Alejandra Valenzuela, BS^49^; Catherine Wandell, BS^21^; Diana Willeke^8^; Karen Williams, BA^3^ and Dilinuer Wubuli, MB^45^

### Partners Scientific Advisory Board (Acknowledgement)

#### Funding

PPMI – a public-private partnership – is funded by the Michael J. Fox Foundation for Parkinson’s Research and funding partners, including 4D Pharma, Abbvie, AcureX, Allergan, Amathus Therapeutics, Aligning Science Across Parkinson’s, AskBio, Avid Radiopharmaceuticals, BIAL, Biogen, Biohaven, BioLegend, BlueRock Therapeutics, Bristol-Myers Squibb, Calico Labs, Celgene, Cerevel Therapeutics, Coave Therapeutics, DaCapo Brainscience, Denali, Edmond J. Safra Foundation, Eli Lilly, Gain Therapeutics, GE HealthCare, Genentech, GSK, Golub Capital, Handl Therapeutics, Insitro, Janssen Neuroscience, Lundbeck, Merck, Meso Scale Discovery, Mission Therapeutics, Neurocrine Biosciences, Pfizer, Piramal, Prevail Therapeutics, Roche, Sanofi, Servier, Sun Pharma Advanced Research Company, Takeda, Teva, UCB, Vanqua Bio, Verily, Voyager Therapeutics, the Weston Family Foundation and Yumanity Therapeutics.

1. Institute for Neurodegenerative Disorders, New Haven, CT
2. University of Luxembourg, Luxembourg
3. Northwestern University, Chicago, IL
4. University of Iowa, Iowa City, IA
5. VectivBio AG
6. The Michael J. Fox Foundation for Parkinson’s Research, New York, NY
7. Innsbruck Medical University, Innsbruck, Austria
8. Paracelsus-Elena Klinik, Kassel, Germany
9. University of California, San Francisco, CA
10. Laboratory of Neuroimaging (LONI), University of Southern California
11. BioRep, Milan, Italy
12. University of Pennsylvania, Philadelphia, PA
13. National Institute on Aging, NIH, Bethesda, MD
14. Mount Sinai Beth Israel, New York, NY
15. Indiana University, Indianapolis, IN
16. Emory University of Medicine, Atlanta, GA
17. Oregon Health and Science University, Portland, OR
18. University of Alabama at Birmingham, Birmingham, AL
19. University of South Florida, Tampa, FL
20. Baylor College of Medicine, Houston, TX
21. University of Washington, Seattle, WA
22. Boston University, Boston, MA
23. University of Rochester, Rochester, NY
24. Banner Research Institute, Sun City, AZ
25. Cleveland Clinic, Cleveland, OH
26. University of Tübingen, Tübingen, Germany
27. University of California, San Diego, CA
28. Johns Hopkins University, Baltimore, MD
29. Imperial College of London, London, UK
30. University of Salerno, Salerno, Italy
31. Parkinson’s Disease and Movement Disorders Center, Boca Raton, FL
32. University of Cincinnati, Cincinnati, OH
33. Hospital Clinic of Barcelona, Barcelona, Spain
34. Hospital Universitario Donostia, San Sebastian, Spain
35. Tel Aviv Sourasky Medical Center, Tel Aviv, Israel
36. St. Olav’s University Hospital, Trondheim, Norway
37. National and Kapodistrian University of Athens, Athens, Greece
38. Columbia University Irving Medical Center, New York, NY
39. Stanford University, Stanford, CA
40. University of Pittsburgh, Pittsburgh, PA
41. Center for Strategy Philanthropy at Milken Institute, Washington D.C.
42. 12, New Brunswick, NJ
43. University of Michigan, Ann Arbor, MI
44. Toronto Western Hospital, Toronto, Canada
45. The Ottawa Hospital, Ottawa, Canada
46. Massachusetts General Hospital, Boston, MA
47. University of Kansas Medical Center, Kansas City, KS
48. University of Southern California, Los Angeles, CA
49. Barrow Neurological Institute, Phoenix, AZ
50. Mayo Clinic Arizona, Scottsdale, AZ
51. University of Colorado, Aurora, CO
52. NYU Langone Medical Center, New York, NY
53. University of Florida, Gainesville, FL
54. Montreal Neurological Institute and Hospital/McGill, Montreal, QC, Canada
55. Cleveland Clinic-Las Vegas Lou Ruvo Center for Brain Health, Las Vegas, NV
56. Clinical Ageing Research Unit, Newcastle, UK
57. John Radcliffe Hospital Oxford and Oxford University, Oxford, UK
58. Universität Lübeck, Luebeck, Germany
59. Radboud University, Nijmegen, Netherlands
60. TransThera Consulting
61. Duke University, Durham, NC
62. Wolfson Institute of Population Health, Queen Mary University of London, UK
63. Philipps-University Marburg, Germany
64. University of Lagos, Nigeria

## Notes

### Competing Interest Statement

H.C. has received research funding from The Michael J. Fox Foundation for Parkinsons Research. D.W. has received research funding or support from The Michael J. Fox Foundation for Parkinsons Research, International Parkinson and Movement Disorder Society, National Institutes of Health, The Parkinsons Foundation, and the US Department of Veterans Affairs; honoraria for consultancy from Boehringer Ingelheim, Cerevel Therapeutics, CHDI Foundation, Citrus Health Group, Medscape, Modality.AI, Roche, Sage Therapeutics, Scion NeuroStim, and Signant Health; and license fee payments from the University of Pennsylvania for the Questionnaire for Impulsive-Compulsive Disorders in Parkinsons Disease (QUIP) and Questionnaire for Impulsive Compulsive Disorders in Parkinsons Disease-Rating Scale (QUIPRS). M.K.Y. has received research funding or support from the Alzheimers Association, the Michael J. Fox Foundation for Parkinsons Research, the Parkinson Foundation, and BlueRock Therapeutics. R.K., A.N., D.E.L, and C.C.G. report research funding to their institution from the Michael J. Fox Foundation for Parkinsons Research.

### Clinical Protocols

https://www.ppmi-info.org/study-design/archive-of-research-docs-and-sops.html

### Funding Statement

This work was supported by The Michael J. Fox Foundation for Parkinsons Research (Michael J. Fox Foundation Write Now Initiative Award).

### Author Declarations

This study used only existing public datasets (PPMI), and therefore did not require review or approval by an Ethics committee/IRB.

### Summary of Updates

Data availability and title has been updated.

## REFERENCES

1. Simuni T, Chahine LM, Poston K, et al. A biological definition of neuronal alpha-synuclein disease: towards an integrated staging system for research. Lancet Neurol 2024;23(2):178–190.

2. Halliday DWR, Mulligan BP, Garrett DD, et al. Mean and variability in functional brain activations differentially predict executive function in older adults: an investigation employing functional near-infrared spectroscopy. Neurophotonics 2018;5(1):011013.

3. Hines LJ, Miller EN, Hinkin CH, et al. Cortical brain atrophy and intra-individual variability in neuropsychological test performance in HIV disease. Brain Imaging Behav 2016;10(3):640–651.

4. Jones RN, Manly J, Glymour MM, Rentz DM, Jefferson AL, Stern Y. Conceptual and measurement challenges in research on cognitive reserve. J Int Neuropsychol Soc 2011;17(4):593–601.

5. Morgan EE, Woods SP, Grant I, Group HIVNRP. Intra-individual neurocognitive variability confers risk of dependence in activities of daily living among HIV-seropositive individuals without HIV-associated neurocognitive disorders. Arch Clin Neuropsychol 2012;27(3):293–303.

6. Stuss DT, Alexander MP. Executive functions and the frontal lobes: a conceptual view. Psychol Res 2000;63(3-4):289–298.

7. Ridgely NC, Woods SP, Webber TA, Mustafa AI, Evans D. Cognitive Intra-individual Variability in the Laboratory Is Associated With Greater Executive Dysfunction in the Daily Lives of Older Adults With HIV. Cogn Behav Neurol 2024;37(1):32–39.

8. Penheiro R, Webber TA, Kiselica AM, Woods SP. Executive Functions are Independently Associated with Cognitive Dispersion in HIV Disease. Arch Clin Neuropsychol 2025;40(2):345–349.

9. Jones JD, Burroughs M, Apodaca M, Bunch J. Greater intraindividual variability in neuropsychological performance predicts cognitive impairment in de novo Parkinson’s disease. Neuropsychology 2020;34(1):24–30.

10. Webber TA, Kiselica AM, Mikula C, Woods SP. Dispersion-based cognitive intra-individual variability in dementia with Lewy bodies. Neuropsychology 2022;36(8):719–729.

11. Combs HL, Henry SK, Webber TA, York MK, Sober JD. “Cognitive functioning and intra-individual variability in essential tremor and Parkinson’s disease: A comparative study”. Clin Neuropsychol 2025:1–21.

12. Davis JJ, Sivaramakrishnan A, Rolin S, Subramanian S. Intra-individual variability in cognitive performance predicts functional decline in Parkinson’s disease. Appl Neuropsychol Adult 2025;32(1):125–132.

13. Jones JD, Valenzuela YG, Uribe C, Bunch J, Kuhn TP. Intraindividual variability in neuropsychological performance predicts longitudinal cortical volume loss in early Parkinson’s disease. Neuropsychology 2022;36(6):513–519.

14. Marek K. The Parkinson’s Progression Markers Inititiative (PPMI) Clinical- Establishing a Deeply Phenotyped PD Cohort AM 4 V.3. protocols.io 2024. 10.17504/protocols.io.n92ldmw6ol5b/v2

15. Marek K, Chowdhury S, Siderowf A, et al. The Parkinson’s progression markers initiative (PPMI) - establishing a PD biomarker cohort. Ann Clin Transl Neurol 2018;5(12):1460–1477.

16. Parkinson Progression Marker I. The Parkinson Progression Marker Initiative (PPMI). Prog Neurobiol 2011;95(4):629–635.

17. Benton AL, Varney NR, Hamsher KD. Visuospatial judgment. A clinical test. Arch Neurol 1978;35(6):364–367.

18. Arango-Lasprilla JC, Rivera D, Garza MT, et al. Hopkins Verbal Learning Test- Revised: Normative data for the Latin American Spanish speaking adult population. NeuroRehabilitation 2015;37(4):699–718.

19. Wechsler D. Wechsler Adult Intelligence Scale- Fourth Edition (WAIS-IV). San Antonio, TX: Pearson, 2008.

20. Heaton R, Miller, SW, Taylor, MJ, Grant, I. Revised comprehensive norms for an expanded Halstead-Reitan battery: Demographically adjusted neuropsychological norms for African American and Caucasian adults. Lutz, FL: Psychological Assessment Resources, 2004.

21. Gladsjo JA, Schuman CC, Evans JD, Peavy GM, Miller SW, Heaton RK. Norms for letter and category fluency: demographic corrections for age, education, and ethnicity. Assessment 1999;6(2):147–178.

22. Smith A. Symbol Digit Modalities Test (SDMT) Manual (Revised). Los Angeles, CA: Western Psychological Services, 1982.

23. Goetz CG, Tilley BC, Shaftman SR, et al. Movement Disorder Society-sponsored revision of the Unified Parkinson’s Disease Rating Scale (MDS-UPDRS): scale presentation and clinimetric testing results. Mov Disord 2008;23(15):2129–2170.

24. Visser M, Marinus J, Stiggelbout AM, Van Hilten JJ. Assessment of autonomic dysfunction in Parkinson’s disease: the SCOPA-AUT. Mov Disord 2004;19(11):1306–1312.

25. Sheikh JI YJ. Geriatric Depression Scale (GDS) Recent evidence and development of a shorter version. Clinical Gerontologist 1986;5:165–173.

26. Weintraub D, Hoops S, Shea JA, et al. Validation of the questionnaire for impulsive-compulsive disorders in Parkinson’s disease. Mov Disord 2009;24(10):1461–1467.

27. Doty RL, Shaman P, Kimmelman CP, Dann MS. University of Pennsylvania Smell Identification Test: a rapid quantitative olfactory function test for the clinic. Laryngoscope 1984;94(2 Pt 1):176–178.

28. Johns MW. A new method for measuring daytime sleepiness: the Epworth sleepiness scale. Sleep 1991;14(6):540–545.

29. Stiasny-Kolster K, Mayer G, Schafer S, Moller JC, Heinzel-Gutenbrunner M, Oertel WH. The REM sleep behavior disorder screening questionnaire--a new diagnostic instrument. Mov Disord 2007;22(16):2386–2393.

30. Nasreddine ZS, Phillips NA, Bedirian V, et al. The Montreal Cognitive Assessment, MoCA: a brief screening tool for mild cognitive impairment. J Am Geriatr Soc 2005;53(4):695–699.

31. Weintraub D, Brumm MC, Kurth R, York MK, Parkinson’s Progression Markers I. Use of Robust Norming to Create a Sensitive Cognitive Summary Score in De Novo Parkinson’s Disease: An Illustrative Example. Mov Disord 2025;40(3):468–477.

32. Costa AS, Dogan I, Schulz JB, Reetz K. Going beyond the mean: Intraindividual variability of cognitive performance in prodromal and early neurodegenerative disorders. Clin Neuropsychol 2019;33(2):369–389.

33. Hilborn JV, Strauss E, Hultsch DF, Hunter MA. Intraindividual variability across cognitive domains: investigation of dispersion levels and performance profiles in older adults. J Clin Exp Neuropsychol 2009;31(4):412–424.

34. Aita SL, Del Bene VA, Knapp DL, et al. Cognitive Intra-individual Variability in Cognitively Healthy APOE epsilon4 Carriers, Mild Cognitive Impairment, and Alzheimer’s Disease: a Meta-analysis. Neuropsychol Rev 2024.

35. Webber TA, Woods SP, Lorkiewicz SA, Yazbeck HW, Schultz ER, Kiselica AM. Cognitive dispersion and its functional relevance in behavioral variant frontotemporal dementia and prodromal behavioral variant frontotemporal dementia. Neuropsychology 2024;38(7):637–652.

36. Tractenberg RE, Pietrzak RH. Intra-individual variability in Alzheimer’s disease and cognitive aging: definitions, context, and effect sizes. PLoS One 2011;6(4):e16973.

